# The Inner Ear and Aging Brain: A Cross-Sectional Study of Vestibular Function and Morphometric Variations in the Entorhinal and Trans-Entorhinal Cortex

**DOI:** 10.1101/2024.06.04.24308371

**Authors:** Claire Vania, Dominic Padova, J. Tilak Ratnanather, Yuri Agrawal

## Abstract

While the vestibular system is widely known for helping to maintain balance, posture, and stable vision, mounting research has also demonstrated its importance in cognition and aging. Recent studies have found that peripheral vestibular loss is significantly associated with deficits in spatial cognitive function in older adults. However, the exact neural pathways through which vestibular loss affects spatial cognitive ability in older adults are unknown. Clinical and subclinical studies have shown that vestibular loss is associated with structural and functional alterations in the spatial cognitive network. Yet, studies on the entorhinal cortex (ERC) and trans-entorhinal cortex (TEC)—two key cortical regions in the vestibular spatial cognitive network—are scarce. This cross-sectional study examines the relationship between vestibular end-organ function and morphometric changes in the ERC and TEC. We used a subset of 103 patients from the Baltimore Longitudinal Study for Aging who had T1-weighted MRI and vestibular physiological data during the same visit to investigate the relationship between age-related vestibular function and the shape of the ERC and TEC. The vestibular end-organ function was evaluated through the cervical vestibular-evoked myogenic potential (cVEMP) test to assess saccular function, the ocular VEMP (oVEMP) test to evaluate the utricular function, and the video head-impulse test (vHIT) to assess semicircular canal function based on vestibular ocular reflex (VOR). Shape alterations were evaluated in terms of surface expansion/contraction relative to the average ERC or TEC shape. Reduced saccular function was associated with surface expansion in the pro-rhinal region of the left ERC, which is hypothesized to be involved in spatial cognitive function, in the intermediate caudal and intermediate superior regions of the right ERC, and the sulcal region of the right TEC. Reduced utricular function was associated with surface contraction in the left lateral TEC and the anterior sulcal and transentorhinal regions of the left ERC, and surface expansion in the lateral region of the left ERC. Reduced canal function is predominantly associated with surface contraction in the pro-rhinal and lateral regions of the right ERC and with posterior sulcal and transentorhinal regions of the right TEC. These end-organ-specific and differential findings highlight the complex relationship between vestibular function and the shapes of the ERC and TEC, key regions that subserve spatial cognitive ability and other crucial cognitive abilities. Future work will determine whether these structural alterations related to vestibular loss may underlie the observed deficits in spatial cognitive ability in older adults and Alzheimer’s disease.

## 1. Introduction

The vestibular system has evolved to sense the motions that arise from head movements and the inertial effects of gravity. Although the vestibular system is widely known for its role in balance, posture, and stable gaze, mounting research demonstrates its importance in cognition and aging (Agrawal *et al*., 2012; Smith *et al*., 2013; Cullen, Fattori and Brichta, 2014). Several recent studies have found that vestibular impairment is significantly associated with cognitive deficits like visuospatial defects in older adults (Bigelow *et al*., 2015; Semenov *et al*., 2016). However, the exact central nervous system pathways through which the loss of peripheral vestibular function affects cognition in older adults remain poorly defined.

Several neuroimaging studies of vestibular function have found structural changes in the vestibular spatial cognitive network, which includes the hippocampus, thalamus, basal ganglia, and the entorhinal cortex (ERC) and trans-entorhinal cortex (TEC) (Hitier, Besnard and Smith, 2014; Yoder, Taube and Besnard, 2014). In a study of 97 healthy, older adults, Padova et al., (2021) found that reduced saccular function was associated with reduced gray matter volumes bilaterally of the basal ganglia and thalamus, and that reduced horizontal canal function was associated with reduced gray matter volume in the left hippocampus (Padova *et al*., 2021). Despite the importance of the ERC and TEC in vestibular and multisensory processing, memory formation and consolidation, spatial memory and navigation, object recognition, and associating objects with their contexts, few studies have linked vestibular function to ERC structure in humans. In a cross-sectional study of 80 healthy older adults, Jacob et al., (2020) found that reduced saccular function was significantly associated with a reduction in the volume of ERC and compression of ERC (Jacob *et al*., 2020). The dearth of ERC and TEC findings could be a consequence of a lack of power due to small sample sizes or inadequate brain mapping methods. The whole-brain voxel-based and volume-based methods that dominate the neuroimaging analysis of vestibular function have known problems with detecting small, non-focal, or non-uniformly distributed effects. Region-of-interest-based shape analysis has emerged to sensitively track structural alterations that such whole-brain voxel-based and volume-based brain mapping methods may miss.

This study investigates the relationship between age-related vestibular end-organ loss and the morphology of the ERC and TEC while adjusting for sex, age, and intracranial volume in 103 older adults from the BLSA using vestibular data and MRI images. This study extends a previous study of the same cohort (Jacob *et al*., 2020) to report new relationships between end-organ function and morphology in the ERC and TEC with new anatomical interpretations. We hypothesized that reduced vestibular function would be associated with surface compressions in the subregions of the ERC and TEC that subserve spatial cognitive ability.

## 2. Methods

### 2.1 Data

To better understand how community-dwelling adults age, the Baltimore Longitudinal Study of Aging (BLSA) was established in 1958 (Shock et al., 1984). We selected a subset of the research participants who underwent brain MRI scans and vestibular testing in the same study visit between 2013 and 2015 and were ≥60 years old. From the total participant cohort, 103 individuals matched the criteria for our study subset. The group’s mean age (± SD) was 76.99 years (± 8.7), and 72 % were males. All participants provided written informed consent, and the BLSA study protocol (03-AG-0325) was approved by the National Institute of Environmental Health Sciences Institutional Review Board.

#### 2.1.1 MRI Testing

The National Institute on Aging’s (NIA) Clinical Research Unit used a 3T Philips Achieva scanner to conduct the MRI scans. Sequences included a T1-weighted volumetric scan magnetization prepared rapid acquisition with gradient echo (MPRAGE; TR = 6.5 ms, TE 3.1 ms, flip angle = 8°, 256 × 256 image matrix, 170 slices, voxel size = 1.0 × 1.0mm, slice thickness = 1.2mm, FOV = 256 × 240mm). T1-weighted volumetric images were collected in the sagittal plane.

#### 2.1.2 Vestibular Testing

Vestibular function testing includes the cVEMP test to assess saccular function, the oVEMP test to assess utricular function, and the video head impulse test (vHIT) which measures VOR gain to assess semicircular canal functioning. A Vestibular-evoked myogenic potential (VEMP) is a short-latency potential that is evoked by activating vestibular receptors with sound or vibration. It is produced by electromyographic impulses that have been modified, either from the inferior oblique muscle for the ocular VEMP (oVEMP) or the sternocleidomastoid muscle for the cervical VEMP (cVEMP). The otolith organs appear to be the source of these reflexes and hence can be used to test the function of otolith organs (Nguyen, Welgampola, and Carey, 2010; Li et al., 2014). A commercial electromyographic system (software version 14.1; Carefusion Synergy, Dublin, OH, USA) was used for recording the VEMP signal. Electromyogram signals were recorded with disposable, self-adhesive, pre-gelled Ag/AgCl electrodes with 40-inch safety lead wires from GN Otometrics (Schaumburg, IL, USA). Electromyogram signals were amplified 2500× and band-pass filtered, 20–2000 Hz for cervical vestibular-evoked myogenic potentials and 3–500 Hz for ocular vestibular-evoked myogenic potentials.

##### a. Cervical VEMP (cVEMP)

Participants laid with upper bodies increased at 30° from horizontal. A non-inverting electrode was placed at the mid-point of the sternocleidomastoid muscle, an inverting electrode was placed on the sternoclavicular junction, and a ground electrode was placed on the manubrium sterni. Bursts of monoaural, 500 Hz, 125 dB sound stimuli were delivered using headphones (VIASYS Healthcare, Madison, WI, USA). The background EMG activity was obtained 10 ms before the start of the sound stimulus, and it was used to normalize the recorded myogenic potentials. In the analyses, the greater cVEMP from either ear was used. By established rules, an absent response was defined as one that fell below a threshold level. For present responses, the higher cVEMP from the left and right sides was used in this analysis.

##### b. Ocular VEMP (oVEMP)

For ocular vestibular-evoked myogenic potential testing, participants laid with upper bodies increased at 30° from horizontal. A non-inverting electrode was placed on the cheek ≈3 mm below the eye, directly beneath the pupil, an inverting electrode was placed 2 cm below the non-inverting electrode, and a ground electrode was placed on the manubrium sterni. Before stimulation, participants were instructed to perform 20° vertical saccades to ensure that symmetrical signals were recorded from both eyes. If signals showed >25% asymmetry, the electrodes were removed, and new ones were applied. Participants were instructed to maintain a 20° up gaze during ocular vestibular-evoked myogenic potential stimulation and recording (Li et al., 2014). The midline of the face was tapped with a reflex hammer at the hairline and about one-third of the space between the inion and nasion. In this investigation, the best response from either ear was used. An absent answer was recorded and repeated for confirmation if the response fell below the predetermined threshold levels (Nguyen, Welgampola, and Carey, 2010; Harun et al., 2016).

##### c. VOR Gain Testing

The horizontal vestibular ocular reflex (VOR) was assessed using the video head impulse test (vHIT). The vHIT was performed in the plane of the right and left horizontal semicircular canals using the EyeSeeCam system (Interacoustics, Eden Prairie, MN, USA). The patient’s head was pitched down 30 degrees to place the horizontal canals in the plane of stimulation, and subjects were asked to fix their gaze on a wall target about 1.5 meters away. The head was moved at a small amplitude (approximately 5–15 degrees) with high velocity (typically 150– 250 degrees per second) horizontally toward the right and left side at least 10 times in both directions. The direction of the head movement was randomized to be unpredictable. The EyeSeeCam system measured eye velocity and head velocity, and a corresponding VOR gain was calculated by dividing the eye velocity by the head velocity. A normal, compensatory VOR gain should equal 1.0. VOR gains less than 0.8, along with compensatory refixation saccades, suggesting a loss of peripheral vestibular function (Harun et al., 2016).

### 2.2 Pipeline

The general outline for shape analysis is shown in Figure 1. A binary segmentation volume is created by segmenting the structure of interest from the brain’s MRI scans. A 3D surface is then obtained by triangulating the binary volume. Shape descriptors were determined by measuring how far each subject’s shape deviated from the population template. Regression is employed in statistical analysis, with shape descriptors as the dependent variable and vestibular factors as the independent variables. The following subsections go into further detail about each phase.

**Figure 1:**
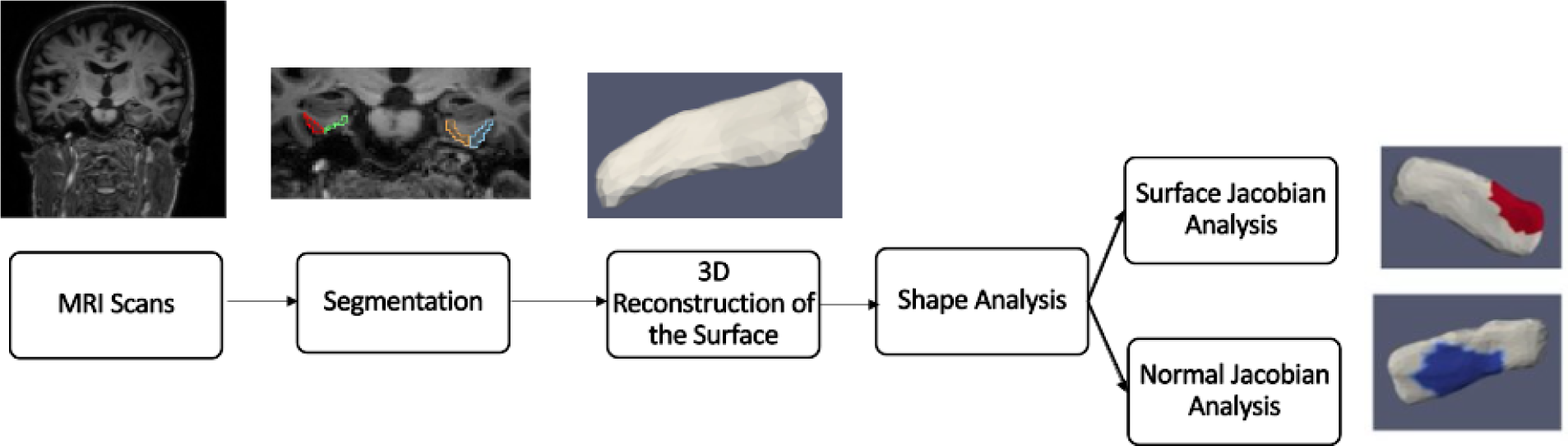
Pipeline Flowchart.

#### 2.2.1 Segmentation and 3D reconstruction of surfaces

T1 scans were automatically segmented by registering them to multiple atlases using Large Deformation Diffeomorphic Metric Mapping (LDDMM) (Beg *et al*., 2005). The parcellation of the scans was based on the Multiple-Atlas Likelihood Fusion (MALF) algorithm (Tang *et al*., 2013, 2015), with 286 defined structures. The parcellation was done through MRICloud, an online neuroinformatics platform that provides tools for automatic brain parcellation and surface registration (https://www.mricloud.org/). Surfaces were created from the binary segmentations using Restricted Delaunay triangulation (Dey and Leekha, 2000; Cohen-Steiner and Morvan, 2003; Cazals and Giesen, 2004). Since the accuracy of the segmentations and the surface reconstruction were crucial to the outcomes, quality control was carried out at every step, and manual editing of segmentations and surface meshes was done when necessary.

#### 2.2.2 Shape Analysis

For shape analysis, the population of surface meshes was rigidly aligned and used to create a ‘mean shape’ called the template. Templates were made separately for each structure’s left and right sides. The generated template is label- and group-blind and serves as a coordinate system for the population average. Each subject surface was registered to this template, first rigidly and then using surface LDDMM. The algorithm computes a smooth invertible mapping of the triangulated surface template onto the target surfaces. MRICloud provides the public with access to this pipeline (Jain *et al*., 2014; Mori *et al*., 2016). The MRICloud diffeomorphic registration pipeline yields two deformation values per vertex: Jacobian determinant of the 3D surface transformation and the surface Jacobian. The Jacobian determinant is a scalar that describes the volume change due to the diffeomorphic change of coordinates. The surface Jacobian is a scalar that describes the ratio between the surface area of the faces attached to a vertex before the transformation (i.e., the surface template) and following it (i.e., the surface template mapped to the target surface). Because surface shape can vary independently in terms of expansion/compression tangent to the surface (i.e. surface area change) and expansion/compression normal to the surface, we compute a third shape descriptor called the normal Jacobian. The normal Jacobian is the ratio between the Jacobian determinant and the surface Jacobian signifies the ratio between the length of an infinitesimal line oriented normally to the surface before and after the transformation. We analyze the surface and normal log-Jacobians independently. A positive (negative) surface log-Jacobian value denotes an expansion (contraction) of the template around that vertex in the direction tangent to the surface to fit the subject. Similarly, a positive (negative) normal Jacobian value denotes an expansion (contraction) of the template around that vertex in the direction normal to the surface to fit the subject. We fit the statistical model to each vertex, thus giving *N* models for each subject, where *N* is the number of vertices in the population template. The null hypothesis is:

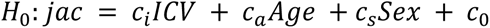

The alternate hypothesis is:

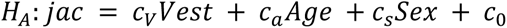

Where *Vest* refers to the vestibular variables investigated (cVEMP, oVEMP, VOR Gain) and *ICV* (Intracranial Volume) refers to the total volume within the skull, including left and right hemispheres, brain stem, cerebellum, and CSF (cerebral spinal fluid); therefore, *ICV* reflects the head size. For each participant, Age in years is a continuous variable and Sex is a binary variable, coded 1 for female and 0 for male. The unknown coefficients {*c*_*V*_, *c*_*i*_, *c*_*a*_, *c*_*s*_} for vestibular variables, *ICV*, age, sex, *c*_0_is the constant term estimated using least-squares. The left and right brain structures are analyzed separately. The schematic of the procedure is shown in Figure 2.

**Figure 2:**
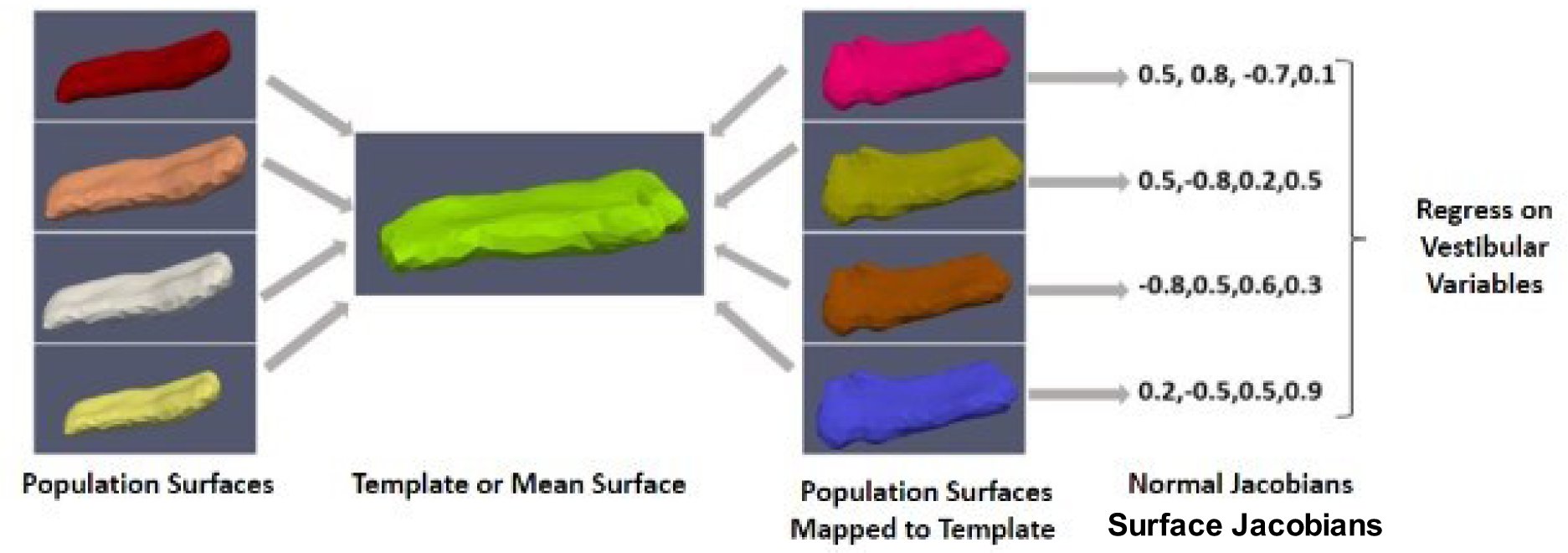
Schematic of Shape Analysis.

We use spectral clustering to partition all vertices (*N* ≈ 400) into *k* (6 to 7) clusters to boost the analytical power and limit the number of models being evaluated simultaneously (Faria *et al*., 2016). Only the shape’s surface geometry is used in this procedure. Each segment’s maximal surface Jacobian is assigned to that cluster, resulting in *k* super-vertices (Jacob *et al*., 2020). In Figure 8, each colored region represents one cluster, thus reducing the number of number of models from *N* (number of vertices, ≈ 400) to *k a* (number of clusters, 6 to 7). Permutation testing is used to calculate significance level or p-value to avoid multiple hypothesis problems (see section 2.2.4). The relative difference was expressed in terms of presence/absence when the categorical form of vestibular variables was used. In the case of continuous form, we report the relative difference with a 1 SD increase in cVEMP, as mentioned above. For completeness, we report our results from our investigation of the relationship between cVEMP and ERC and TEC surface Jacobians which was also investigated in a previous study of the same cohort (Jacob *et al*., 2020).

**Figure 4:**
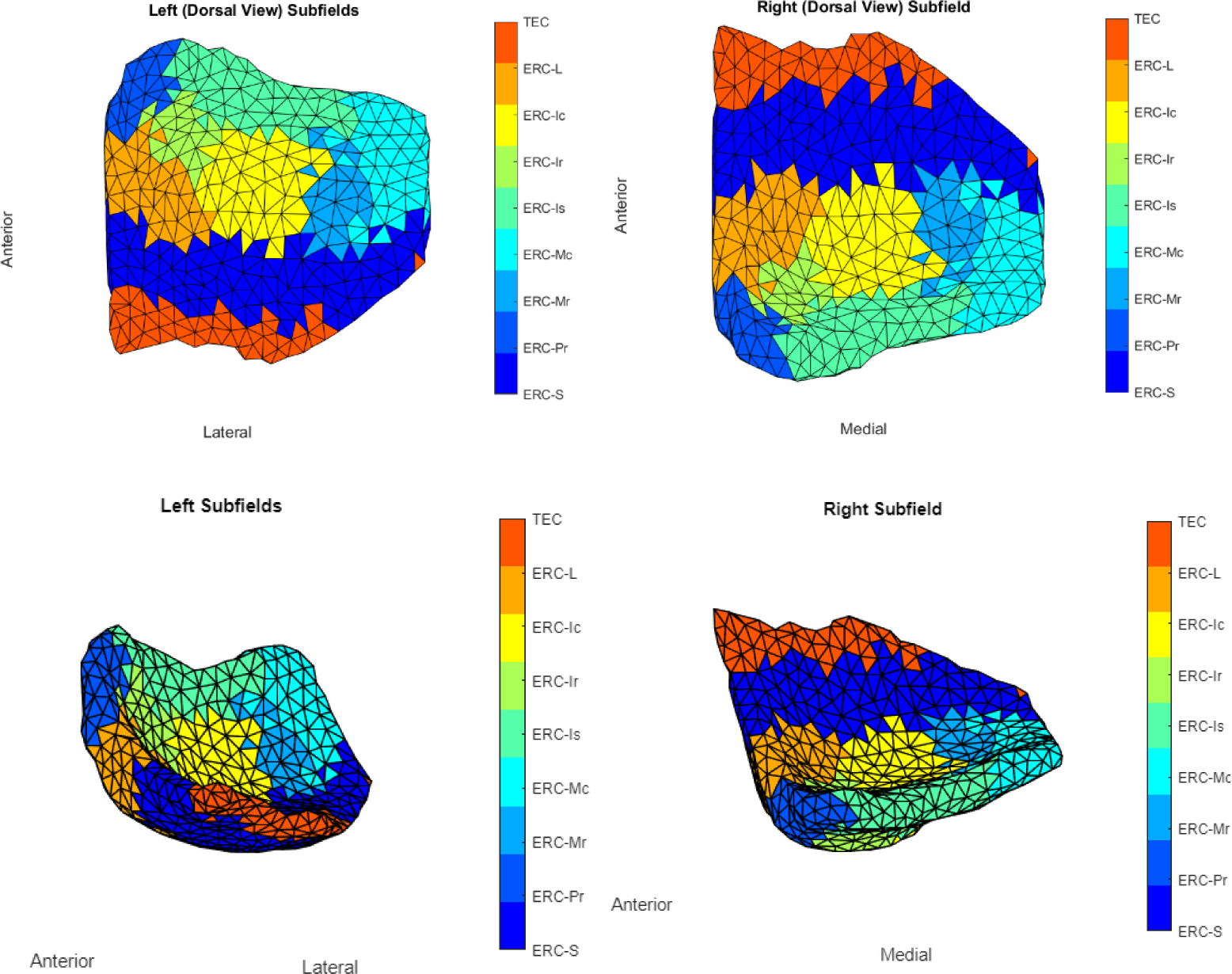
View of ERC Overlayed with Krimer Partitions

**Figure 5:**
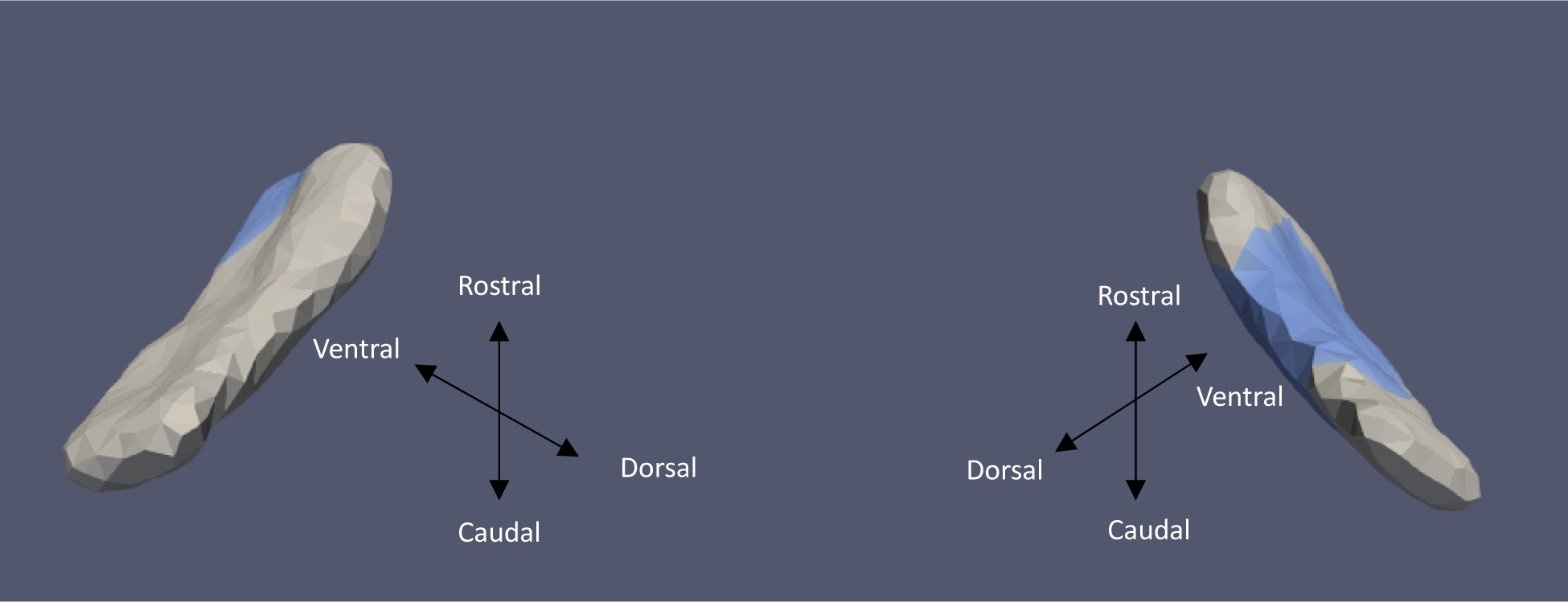
ERC Template showing significant results for the clustered Left and Right sides of the ERC (view from the caudal end, upside-down) for categorical cVEMP.

**Figure 6:**
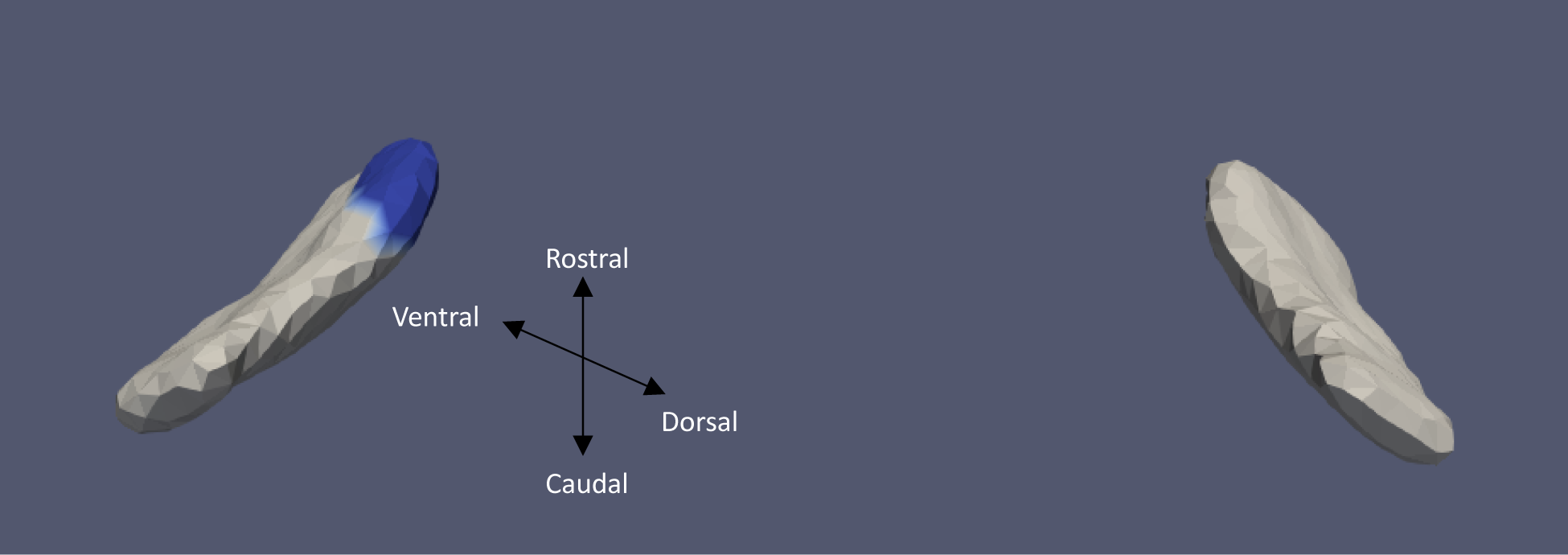
ERC Template showing significant results for the clustered Left and Right sides of the ERC (view from the caudal end, upside-down) for categorical oVEMP.

**Figure 7:**
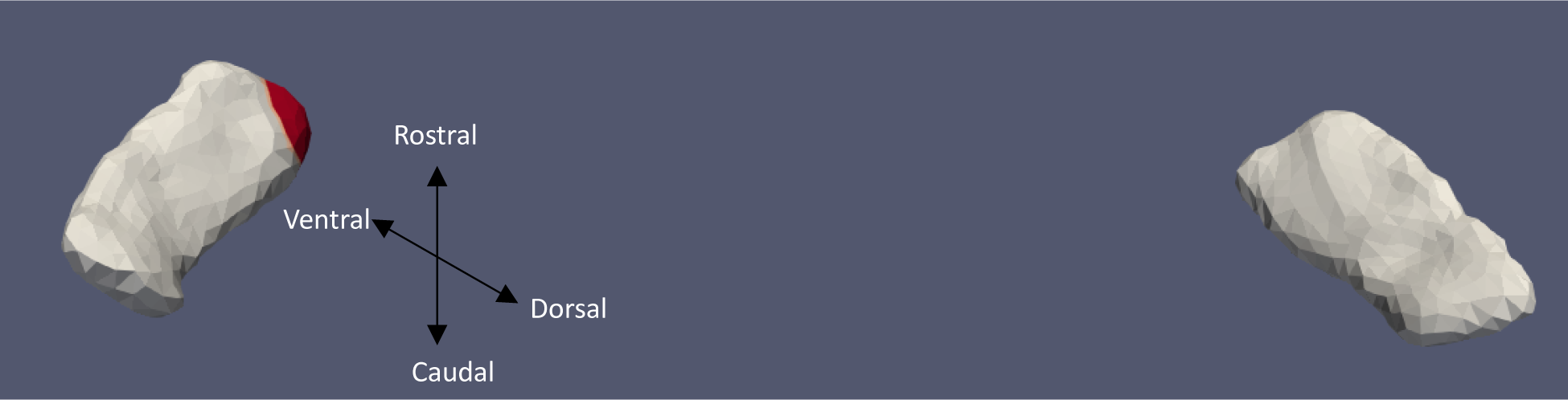
TEC Template showing significant results for the clustered Left and Right sides of the TEC (view from the caudal end, upside-down) for continuous oVEMP.

**Figure 8:**
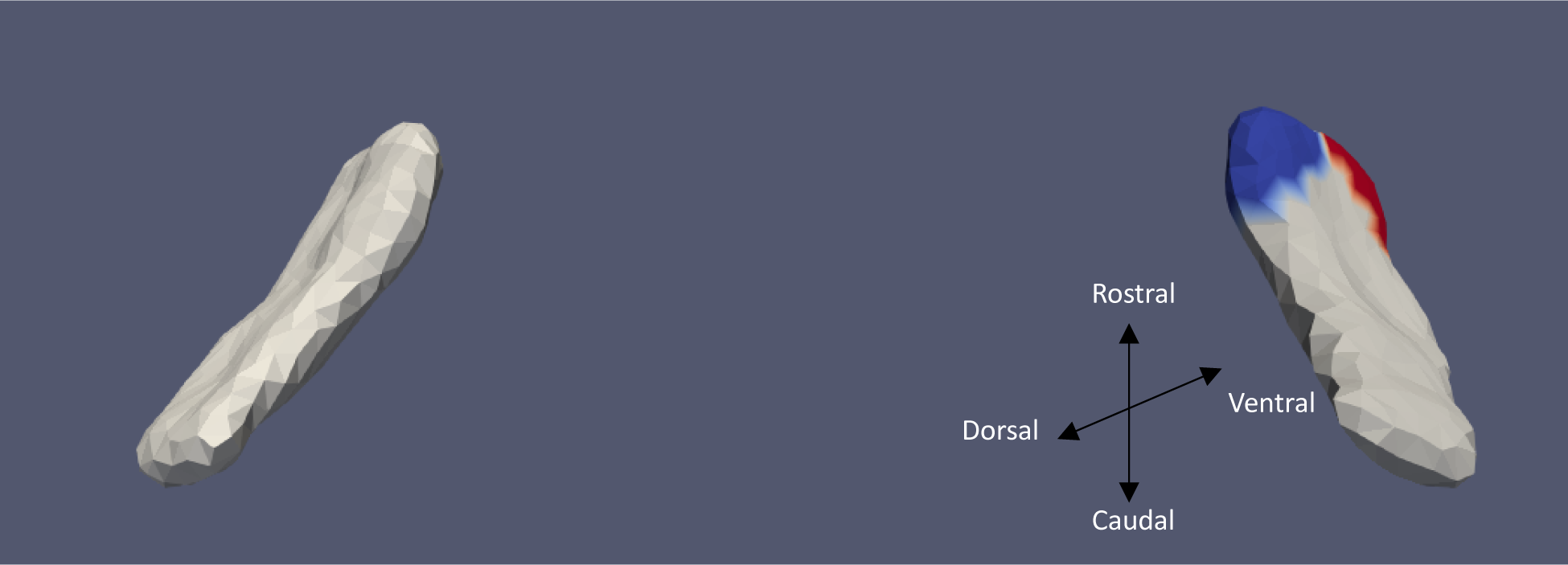
ERC Template showing significant results for the clustered Left and Right sides of the ERC (view from the caudal end, upside-down) for categorical VOR.

**Figure 9:**
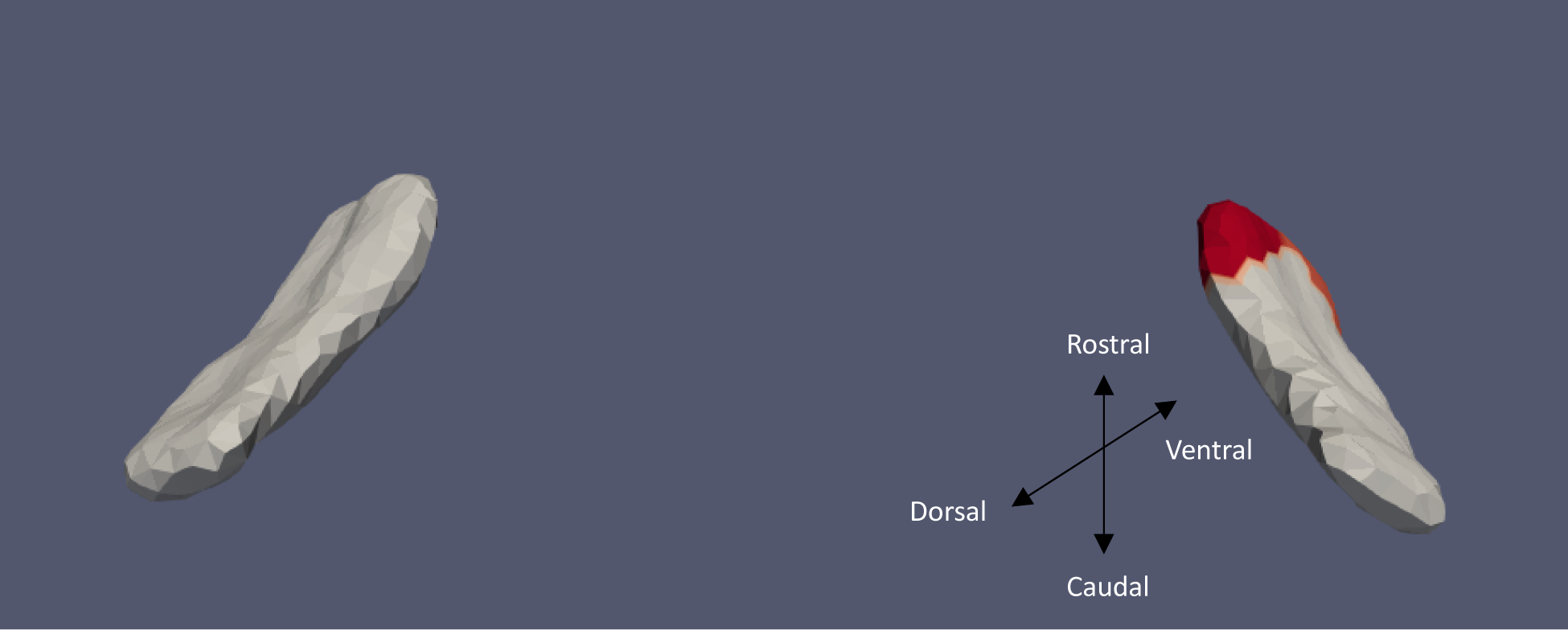
ERC Template showing significant results for the clustered Left and Right sides of the ERC (view from the caudal end, upside-down) for continuous VOR.

#### 2.2.3 Multiple Hypothesis Testing

The problem of multiple hypothesis testing arises from fitting a linear model to each cluster. The probability of making at least one Type I error is significantly higher than alpha if the same procedure is applied to several hypotheses tested simultaneously, particularly when the number of hypotheses is large. Here we use permutation testing to control the Family Wise Error Rate (FWER). Permutation tests build an empirical estimate of the distribution of the test statistic under the null hypothesis by permuting/resampling data. The test statistic from the real data is then compared to this distribution, and the p-value is the fraction of the population greater than the real test statistic. Here, the test statistic was chosen as a ratio of maximum square errors in the null hypothesis to maximum square error in the alternate hypothesis.

### 2.3 High Field Atlasing

We used a high field strength (11T) MRI to label structures of interest on the MRI images used in this study as *a posteriori*. To help us define boundaries for our measurements of the ERC, we used the partitions established by Krimer et al (Krimer *et al*., 1997). The Krimer partition, as it has been referred to throughout this study, divides the ERC into nine sub-regions. Established protocols were used to construct the lateral to medial coordinates for the 9 subregions (Krimer *et al*., 1997; Insausti *et al*., 1998; Miller *et al*., 2015). This study defines the following 7 Krimer regions together as the ERC: 1. Intermediate Superior (ERC-Is), 2. Intermediate Rostral (ERC-Ir), 3. Intermediate Caudal (ERC-Ic), 4. Pro-rhinal (ERC-Pr), 5. Medial Rostral (ERC-Mr), 6. Medial Caudal (ERC-Mc), and 7. Lateral (ERC-L). This definition corresponds well with how the ERC is typically defined for most MRI studies. We represent the Krimer sulcal (ERC-S) and trans entorhinal regions together as the TEC. To assign Krimer labels to the population template, we first rigidly aligned the population template and the high-field atlas, and then we used surface LDDMM to map the population template onto the high-field atlas. Krimer labels were transferred to the population template vertices using the deformation field followed by a nearest-neighbor label assignment. Fig 4 shows the ERC population template with Krimer subfields. In a prior study, Kulason et al., 2020, highlighted the discrepancies stemming from different nomenclature about the TEC and ERC. Their comparative examination of four atlases provided insight into these disparities with Figure 4 in the paper serving as a focal point for reference.

## 3. Results

### 3.1 Demographic Data

We had 80, 83, and 91 participants who had their cVEMP, oVEMP, and VOR measured, respectively, and MRI scans taken during the same visit. Mean cVEMP, oVEMP, and VOR amplitudes and the mean age for each cohort are shown in Table 1.72% of participants in the cVEMP cohort and 77% of participants in the oVEMP cohort had response present in either ear. 8% of participants had impaired canal function. Table 2 shows the percentage of continuous and discontinuous sulcus in each cohort. Several variants of continuous collateral sulcus (CoS) exist. The first variant, Type I CoS, is characterized by a deep, continuous sulcus where the rhinal sulcus shares a sulcal bed with the collateral sulcus proper (Ding and Van Hoesen, 2010). This pattern has also been identified as Type II/Type III rhinal sulcus (Huntgeburth and Petrides, 2012). The second variant, termed Type IIa CoS, exhibits a discontinuous CoS where the collateral sulcus proper starts posterior to the GI (Ding and Van Hoesen, 2010). Additionally, the Type I rhinal sulcus, aligns with Type IIa CoS (Huntgeburth and Petrides, 2012). Finally, there is the Type IIb CoS, distinguished by a discontinuous pattern where the collateral sulcus proper begins anterior to the GI (Ding and Van Hoesen, 2010). Despite the differences in morphological sub-type of the collateral sulcus, we include all ERC and TEC surfaces in our analysis to maintain our sample size.

**Table 1:** Characteristics of Study Sample (N=103).

| Characteristic | Mean (SD) | N (%) |
| --- | --- | --- |
| Age | 76.99 (8.64) |  |
| Intracranial Volume (mm<sup>3</sup>) | 1231.1 (118.38) |  |
| **Gender** |  |  |
| Male |  | 74 (72) |
| Female |  | 29 (28) |
| **Race** |  |  |
| White |  | 75 (73) |
| Black |  | 22 (21) |
| Other |  | 6 (6) |
| **Response Present in Either Ear** |  |  |
| cVEMP |  | 72 |
| oVEMP |  | 77 |
| **VOR Gain** | 0.98 (0.15) | 8 |

**Table 2:**
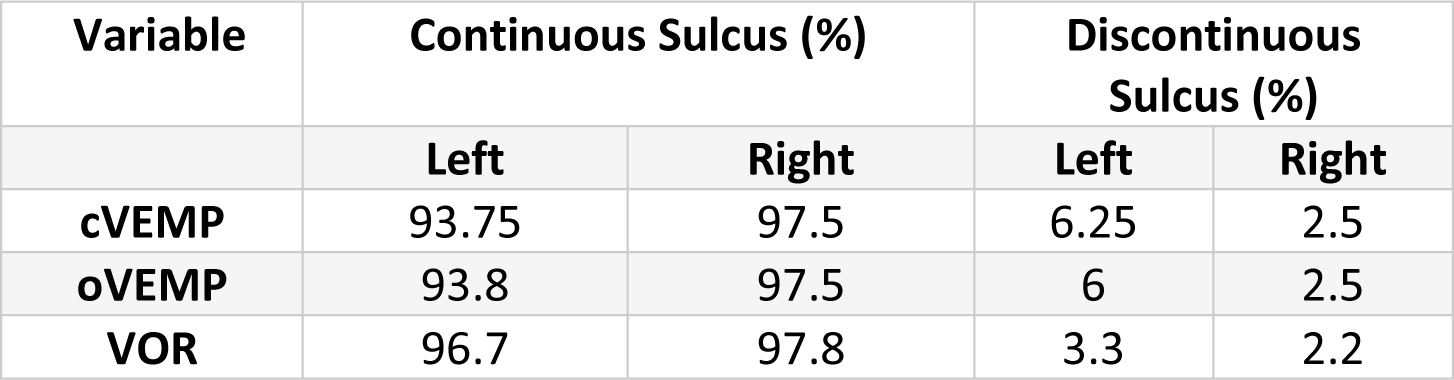
Collateral sulcus distribution.

### 3.2 Results of the Surface Jacobian Analysis

Tables 3 and 4 summarize the significant results of the surface Jacobian analysis for the ERC and the TEC, respectively. In the tables, multiple coefficients denote multiple significant clusters. Figures 5, 6, 8, 9, and 7 illustrate the spatial arrangement of the vestibular effects concerning the surface Jacobian on the ERC template and TEC template, respectively. In the figures, the colors red and blue, respectively, represent expansion and compression relative to the population template.

**Table 3:** Summarizing the significant results of the surface Jacobian analysis for the ERC. A positive (negative) coefficient *c*_*V*_ indicates surface expansion (contraction) in the direction tangent to the surface.

| Form of Vestibular Variable | Side | Coefficient ($c_V$) | % change | Krimer Region | p_perm |
| --- | --- | --- | --- | --- | --- |
| **Categorical cVEMP** | Left | -0.0840 | -0.08 | Pro-rhinal | 0.01 |
|  | Right | -0.108 | -0.1 | Intermediate Caudal & Intermediate Superior | 0.03 |
| **Categorical - oVEMP** | Left | -0.07 | -0.07 | Lateral | 0.03 |
| **Categorical VOR** | Right | 0.25, -0.03 | 0.2, - 0.3 | Pro-rhinal, Lateral | 0.01 |
| **Continuous VOR** | Right | 0.25, 0.27 | 0.25,0.27 | Lateral, Pro-rhinal | 0.008 |

**Table 4:**
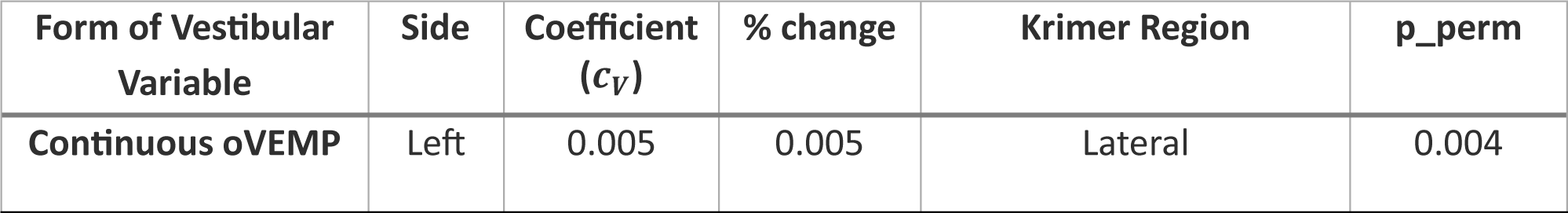
Summarizing the significant results of the surface Jacobian analysis for the TEC. A positive (negative) coefficient *c*_*V*_ indicates surface expansion (contraction) in the direction tangent to the surface.

Categorical cVEMP showed a significant relationship with the left (p ≈ 0.01) and right (p ≈ 0.03) sides of the ERC and compression of, respectively, 0.08% in the anterior medial region and 0.1% in the inferior medial region. The TEC and cVEMP did not exhibit any significant correlation, and continuous cVEMP did not indicate any significance with the ERC. Categorical oVEMP demonstrated no link with the right side of the ERC and a substantial relationship with the left ERC (p ≈ 0.03) with compression of 0.07% in the anterior lateral region. Continuous oVEMP was unrelated to the ERC. The TEC didn’t exhibit any relationship with categorical oVEMP. Continuous oVEMP with compression of 0.05% in the anterior lateral region significantly related to the left side (p ≈ 0.04) of TEC. There was no relationship between the right TEC and continuous oVEMP. With an expansion of 0.25% in the anterior medial region and a compression of 0.03% in the anterior lateral region, categorical VOR demonstrated an association with the right ERC (p ≈ 0.01). With an expansion of 0.2% in the anterior region, right ERC (p ≈ 0.008) also correlated with continuous VOR. There was no correlation between left ERC and VOR. Moreover, there was no association between the TEC and VOR gain.

### 3.3 Results of the Normal Jacobian Analysis

Tables 5 and 6 summarize the significant results of the normal Jacobian analysis for the ERC and the TEC, respectively. As in the previous section, analyses were conducted using continuous and categorical formats for all vestibular variables (cVEMP, oVEMP, and mean VOR gain). Figures 10, 13, 14, 11, 12, and 15 illustrate the spatial arrangement of the vestibular effects concerning the normal Jacobian on the ERC template and TEC template, respectively. In the figures, the colors red and blue, respectively, represent normal surface compression or normal surface expansion relative to the mean template.

**Table 5:** Summarizing the significant results of the normal Jacobian analysis for the ERC. A positive (negative) coefficient *c*_*V*_ indicates surface expansion (contraction) in the direction normal to the surface.

| Form of Vestibular Variable | Side | Coefficient ($c_V$) | % change | Krimer Region | p_perm |
| --- | --- | --- | --- | --- | --- |
| Categorical - cVEMP | Left | - 0.04 | - 0.04 | Pro-rhinal | 0.009 |
| Categorical - VOR | Right | 0.01 | 0.014 | Pro-rhinal | 0.04 |
| Continuous - VOR | Right | 0.13 | 0.12 | Pro-rhinal | 0.03 |

**Table 6:** Summarizing the significant results of the normal Jacobian analysis for the TEC. A positive (negative) coefficient *c*_*V*_ indicates surface expansion (contraction) in the direction normal to the surface.

| Form of Vestibular Variable | Side | Coefficient ($c_V$) | % change | Krimer Region | p_perm |
| --- | --- | --- | --- | --- | --- |
| Categorical - cVEMP | Right | -0.18 | - 0.18 | Sulcal | 0.04 |
| Continuous - oVEMP | Left | 0.004 | 0.004 | Anterior Sulcal and Transentorhinal | 0.02 |
| Categorical - VOR | Right | -0.13 | - 0.12 | Posterior Sulcal & Transentorhinal | 0.04 |

**Figure 10:**
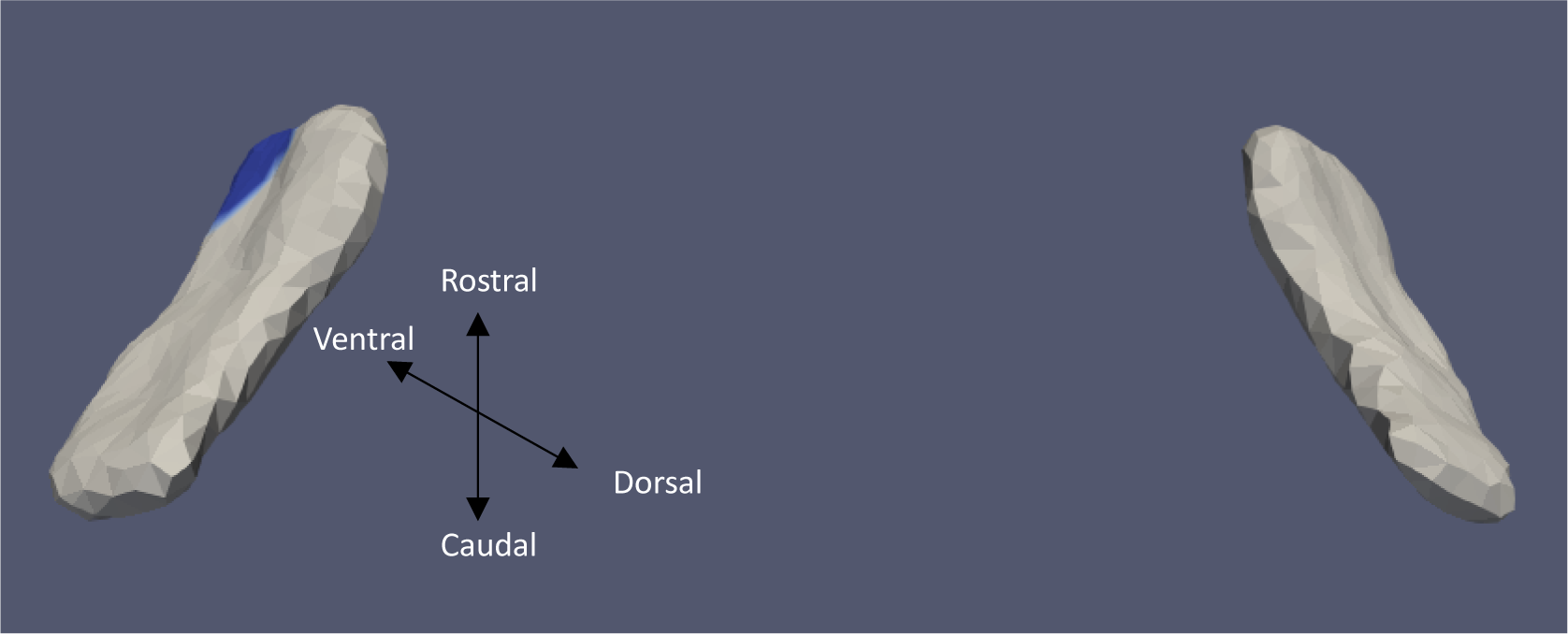
ERC Template showing significant results for the clustered Left and Right sides of the ERC (view from the caudal end, upside-down) for categorical cVEMP.

**Figure 11:**
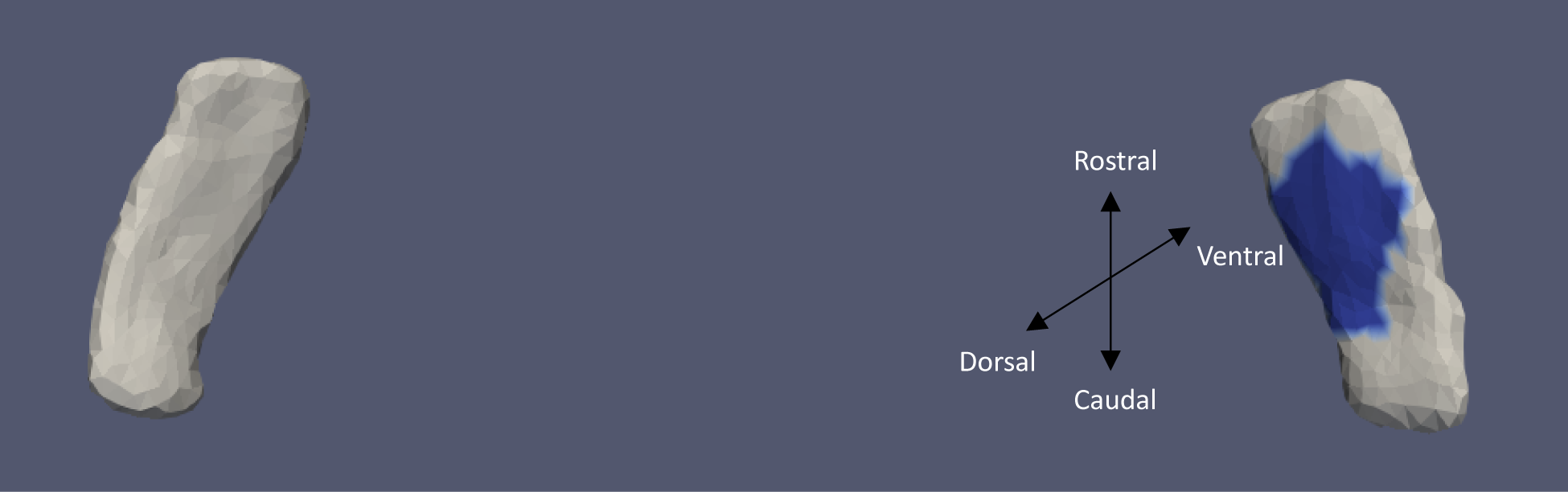
TEC Template showing significant results for the clustered Left and Right sides of the TEC (view from the caudal end, upside-down) for categorical cVEMP.

**Figure 12:**
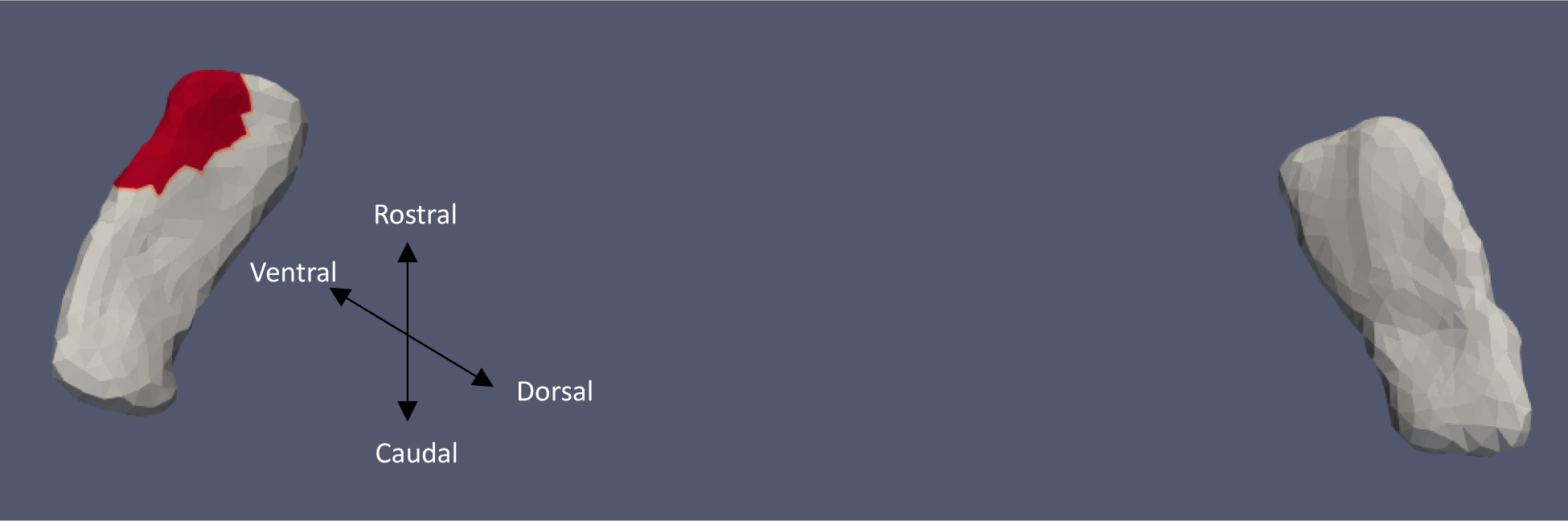
TEC Template showing significant results for the clustered Left and Right sides of the TEC (view from the caudal end, upside-down) for continuous oVEMP.

**Figure 13:**
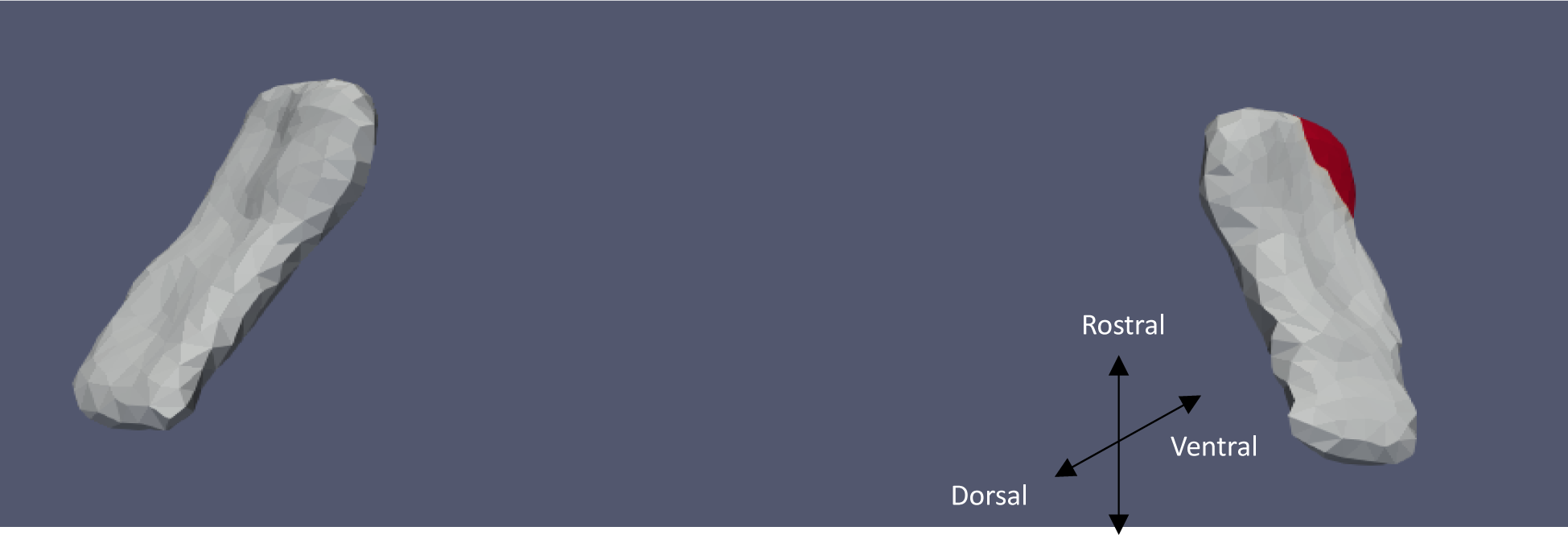
ERC Template showing significant results for the clustered Left and Right sides of the ERC (view from the caudal end, upside-down) for categorical VOR.

**Figure 14:**
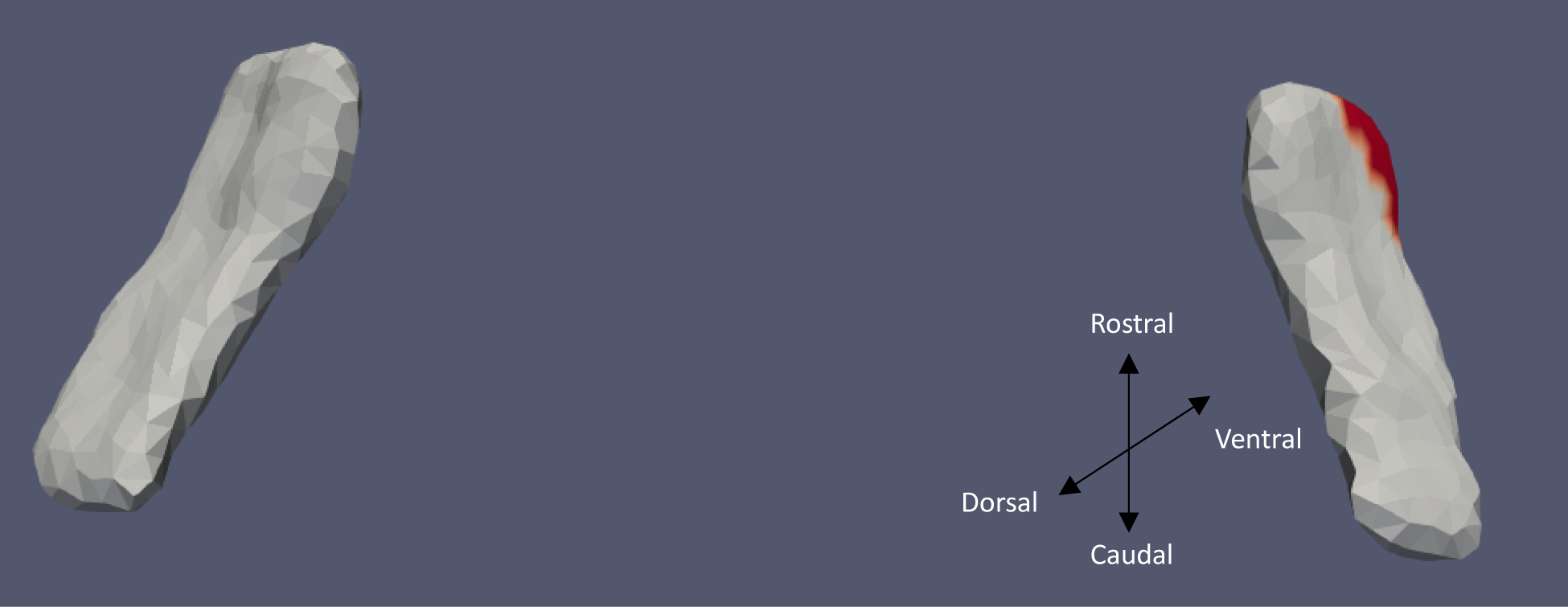
ERC Template showing significant results for the clustered Left and Right sides of the ERC (view from the caudal end, upside-down) for continuous VOR.

**Figure 15:**
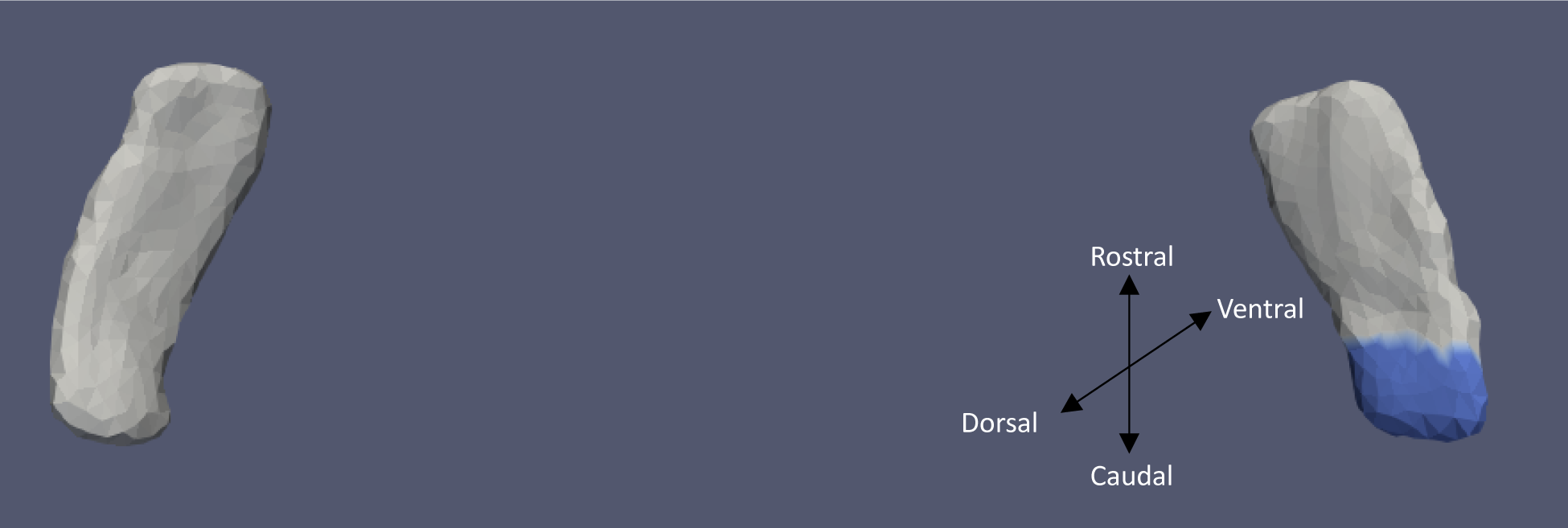
TEC Template showing significant results for the clustered Left and Right sides of the TEC (view from the caudal end, upside-down) for categorical VOR.

Categorical cVEMP showed a significant relationship with the left side of ERC (p ≈ 0.009) and the right side of TEC (p ≈ 0.04) and normal compression of 0.04% in the anterior medial region and 0.18% in the inferior ventral lateral region, respectively. Continuous cVEMP showed no significance with TEC or ERC. Continuous oVEMP showed significance with left TEC (p ≈ 0.02) with a normal expansion of 0.004% in the anterior lateral region. No significance was seen between ERC and right TEC. Categorical VOR and Continuous VOR show significance with the right side of ERC, with a normal expansion of 0.01% (p ≈ 0.04) in the anterior medial region and 0.12% (p ≈ 0.03) also in the anterior medial region, respectively. No significance between left ERC and VOR is seen. Categorical VOR shows a significant association with the right side of TEC, with a normal compression of 0.12% (p ≈ 0.04) in the posterior region. No association was seen between continuous VOR and TEC.

## 4 Conclusion

### 4.1 Discussions

In this study of healthy older adults, we analyzed the relationship between age-related vestibular end-organ loss and the shapes of the ERC and TEC—two cortical regions that putatively receive vestibular input and subserve spatial cognitive function. Our work significantly extends a previous study of the same cohort (Jacob *et al*., 2020) which examined the relationship between saccular function as measured by cVEMP and ERC and TEC surface shape as measured by the surface Jacobian. We found cVEMP to be significantly related to the shape of the ERC, showing a relationship between reduced vestibular function and compression of the ERC. This was also reflected in the normal Jacobian relationship, which shows surface compression with reduced vestibular function. oVEMP and the shape of the ERC also displayed a similar correlation. The compressions are seen in the prorhinal, intermediate superior, and intermediate caudal Krimer regions, which correspond to anterior medial portion of the ERC manual structural labels based on cortical folding, as shown in (Kulason *et al*., 2020). Furthermore, compressions were observed in the lateral Krimer region, corresponding to the anterior lateral portions of the ERC manual structural labels (Kulason *et al*., 2020). The lateral entorhinal cortex provides one of the two major input pathways to the hippocampus and has been suggested to process associative recognition memory (Huntgeburth and Petrides, 2012). The dysfunction of the medial ERC results in spatial memory deficits observed in both animals and humans. Moreover, it is linked with compromised spatial cognition in individuals with Alzheimer’s disease (Malone *et al*., 2024). The presence of grid cells, head direction cells, speed-modulated cells, and border/boundary cells in the medial entorhinal cortex further cements the idea of it being involved in the spatial processing system (Sargolini *et al*., 2006; Solstad *et al*., 2008). These results align with a growing body of research in both humans and animals showing vestibular modulation of entorhinal function (Jacob et al., 2014; Rancz et al., 2015; Simon et al., 2015; Leong et al., 2023). Additionally, it has been demonstrated that bilateral vestibular deactivation or lesions in rats cause theta rhythm, and pattern of brain waves that is crucial for the coordination of place cells, to be disrupted in CA1 and the ERC (Russell *et al*., 2006; Baek *et al*., 2010). Interestingly, with VOR gain, we see an expansion of the ERC. This is also reflected in the normal Jacobian relationship, which shows a positive correlation with deviation of VOR gain from normal. Specifically, the more abnormal the average VOR, the more the increase in normal Jacobian. This can be explained by the ERC’s interconnection with the hippocampus and hippocampus receiving projections from the visual centers of the brain. Jacob et al. 2020, observed an expansion in the local surface of the hippocampus with reduced vestibular function (Jacob *et al*., 2020). The ERC projects strongly to CA1 and CA3 of the hippocampus (Brun *et al*., 2002). Signals from the semicircular canal and otolith organ converge on the medial vestibular nuclei. This vestibular information contributes to the head direction signal, which ascends to the anterior dorsal thalamus and then to the post subiculum which projects to the entorhinal cortex (Yoder, Taube, and Besnard, 2014). Although we see a connection between vestibular inputs, ERC, and hippocampus, further investigation would be required to understand why there is an increase in surface area and normal Jacobian in these regions to poorer vestibular function. Disparities have also been noted in children with ADHD, where variations in basal ganglia volume change sometimes contradicted the direction of local shape alterations (Seymour *et al*., 2017; Tang *et al*., 2019).

For the TEC, we see a similar relationship between normal Jacobian and cVEMP as the ERC; there is a normal surface compression with reduced vestibular function. VOR and oVEMP significantly correlate with the TEC normal Jacobian and surface Jacobian, respectively, but the correlations are opposite to what we get for ERC. With VOR, we see a decrease in normal Jacobian with a more abnormal average VOR. Reduced oVEMP shows a tangential expansion in the shape of the TEC. The results of the TEC, like the ERC, can be attributed to the fact that the TEC, which contains Brodmann area 35, projects strongly to ERC (Burwell, 2000). The decrease in VOR normal Jacobian with abnormal VOR aligns with the proposition that the TEC is deemed to lie at the apex of a hierarchy of processing in the ventral visual stream (Bussey and Saksida, 2007; Ferko *et al*., 2022). We observe these changes in the perirhinal cortex comprising sulcal and transentorhinal regions of the Krimer atlas. The perirhinal cortex plays a major role in object recognition and gives out direct projections to the lateral entorhinal cortex further emphasizing the relationship proposed in our findings.

### 4.2 Limitations

A fundamental limitation of this study is that the results critically depend on the segmentation quality. Due to their blurry and ambiguous grey matter-white matter boundaries the boundary along the meninges, and their proximity to the oculomotor nerve, cortical structures like the entorhinal and trans-entorhinal cortex are notoriously challenging to segment automatically. Furthermore, the diversity of cortical shapes and structures leads to high interobserver variability, necessitating manual segmentation by an expert. The quality of manual segmentation also varies from expert to expert. Another limitation comes from spectral clustering: a strong signal region could be averaged over in a cluster to increase power, but it could also be divided into several clusters to decrease power. Additionally, using a low number of clusters reduces the resolution, restricting our ability to localize to areas smaller than a cluster size. Whether our findings can be interpreted in terms of shape or thickness atrophy is limited. Because the surface Jacobian measuring compression and expansion reflects changes on the surface of the structure, it does not describe changes inside the structure. This limits the ability to claim whole-shape atrophy. Furthermore, the normal Jacobian differs in thickness. Whereas the normal Jacobian describes the scaling factor by which a deformed surface expanded/contracted normally to the surface at a point, thickness is defined as the lengths of the normal lines spanning two points, each on opposing surface sides (e.g. the gray-white matter surface and the gray-pial surface). A change in thickness may not imply a change in normal Jacobian, and similarly vice versa. Future histological studies would be needed to corroborate if the shape changes reported here indicate shape or thickness atrophy.

The best value from either ear is used for cVEMP and oVEMP. As a result, any side specificity is lost. This could also skew the results because if we do not account for the measurement side, we may lose any contralateral or ipsilateral dependence of shape on vestibular function. In addition, we lack knowledge of the subjects’ potential confounding variables, such as handedness, medication, etc. As a result, the population may develop biases, which would then be reflected in the biased template. Another limitation is that the shape analysis template is based on a specific sample. The BLSA template (and resulting analyses) may not be entirely generalizable for the larger adult population to the extent that BLSA participants may differ from the general population of that same age range. Furthermore, it is unknown whether younger people with impaired vestibular function experience the same changes in brain structures that we saw in this population. Further confirmation of our analyses in other populations will be needed to confirm the robustness of our findings.

### 4.3 Future Work

The relationships between age-related vestibular function and the shapes of the ERC and TEC were examined in this study. Even though a cross-sectional study of this kind helps observe relationships between various factors and structures, it does not offer conclusive details about cause-and-effect relationships. As a next step, the hypothesis that vestibular loss precedes structural atrophy can be tested in a longitudinal study. To investigate simultaneous or bidirectional relationships between vestibular function and structure, longitudinal data can be used in conjunction with statistical techniques like structural equation modeling. Additionally, changepoint analysis can be applied to longitudinal data to determine when non-linearities occur in trajectories of structural change. Furthermore, advanced analysis techniques such as network modeling can be used to evaluate how vestibular impairment in older adults may lead to a network of changes that co-occur or in characteristic sequences and how these changes relate to cognitive phenotype. These studies together may provide insights into the role of the peripheral vestibular system in spatial cognitive function in older adults.

## Acknowledgments

This work was supported in part by the National Institute on Aging [grant number R01 AG057667], National Institute on Deafness and Other Communication Disorders [grant number R03 DC015583], and National Institute of Biomedical Imaging and Bioengineering [grant number P41-EB031771] and by the Intramural Research Program, National Institute on Aging, National Institutes of Health.

## Data Availability Statement

Data from the BLSA are available on request by proposal submission through the BLSA website (blsa.nih.gov). All requests are reviewed by the BLSA Data Sharing Proposal Review Committee and are also subject to approval from the NIH institutional review board.

